# The Clinical and Cellular Landscape of DKK1 in Osteosarcoma

**DOI:** 10.64898/2026.11.03.26367177

**Authors:** Esteban Uceda Arias-Stella, Laurel Kastner, Noah Copperman, Genesis Then, Jonathan Karlin, Aakash Mahant Mahant, Masanori Hayashi, Michael Isakoff, Jessica Crimella, Michelle Ewart, Rebeca San Martin, Arthur Berg, Carl Gregory, David M. Loeb

## Abstract

**Background:** Dickkopf-1 (DKK1) is a secreted protein that acts as a Wnt antagonist and is implicated in osteosarcoma progression. Elevated circulating DKK1 levels have been described at OS diagnosis, but its circulating dynamics, prognostic relevance, and cellular distribution remain incompletely characterized.

**Methods:** We analysed longitudinal plasma DKK1 in 34 patients using mixed-effects models, evaluated *DKK1* expression and survival in two independent transcriptomic cohorts, and examined cellular and tissue distribution using single-cell transcriptomics and immunohistochemistry. Cultured mesenchymal stem cells were assessed as potential nonmalignant sources.

**Results:** Circulating DKK1 declined by 12.4% per 30-day period during treatment, without significant changes detected around primary tumour resection or relapse. Metastatic presentation and poor histologic response were associated with higher estimated DKK1 levels. Higher tumour *DKK1* expression was associated with worse overall and metastasis-free survival (pooled HRs per SD increase: 1.35 [95% CI 1.02–1.78] and 1.48 [1.06–2.07], respectively). *DKK1* transcripts were predominantly detected in malignant cells across multiple subpopulations and protein staining was enriched in osteoid and matrix-rich regions of primary tumours.

**Conclusions:** Tumour DKK1 expression is associated with adverse outcomes, while circulating concentration decline over treatment time. These findings support further prognostic evaluation and investigation of the associations among DKK1 expression and production, tissue localisation, and circulating levels.

## Introduction

One of the central challenges in the treatment of osteosarcoma (OS) is metastatic progression. Although the combination of surgery and multi-agent chemotherapy results in long-term survival in approximately 70% of patients with localized disease, outcomes remain poor for patients with metastatic or recurrent OS [1]. Attempts to intensify conventional chemotherapy have failed to improve outcomes, while many clinical trials investigating systemic therapies for recurrent disease have likewise yielded limited benefits [2, 3]. These limitations highlight the need to identify molecular pathways involved in metastatic progression, particularly those that may be therapeutically targeted or evaluated through circulating biomarkers.

Dickkopf-1 (DKK1) is a secreted protein that acts as an antagonist of canonical Wnt signalling through interaction with the LRP5/6 and Kremen-1/2 coreceptors [4, 5]. In addition, DKK1 can interact with CKAP4, activating the AKT signalling pathway [6]. During normal bone development, DKK1 blocks the differentiation and maturation of osteoprogenitors by suppressing Wnt signalling and can shift lineage commitment in mesenchymal stem cells (MSCs) to adipogenesis [7, 8]. DKK1 is often secreted by osteoblasts, resulting in negative regulation of bone formation [9].

In OS, DKK1 has been associated with impaired osteogenic differentiation, increased tumour growth and bone destruction, and activation of ALDH activity [10, 11]. We have previously shown that DKK1-directed interventions reduce metastatic progression in OS models [11-13]. In addition, a recent analysis combining bulk and single-cell transcriptomic data nominated DKK1 as a candidate prognostic factor [14]. Together, these findings implicate DKK1 in tumour progression and suggest that its effects on osteogenic differentiation and bone homeostasis may contribute to interactions between tumour cells and their surrounding microenvironment.

As a secreted protein, DKK1 is also detectable in circulation. Newly diagnosed OS presents with elevated serum DKK1 compared to healthy controls, suggesting potential utility as a biomarker [15]. However, the interpretation of serum DKK1 is complicated by its potential sources, which can include both normal and malignant cells. Activated platelets secrete DKK1 [16], and experimental work in breast cancer has identified contributions from tumoral and stromal compartments, as well as normal bone [17]. Additionally, in breast cancer, serum DKK1 levels are elevated compared to healthy controls, and even more increased in patients with bone metastases, suggesting the bone microenvironment is especially relevant as a potential source of circulating DKK1 [18]. This provides a rationale for evaluating circulating DKK1 in OS patients longitudinally to understand whether circulating DKK1 concentrations change during treatment, reflect disease burden, or correlate with clinical characteristics.

In this study, we quantified plasma DKK1 concentrations during OS treatment and follow-up and assessed associations with metastatic presentation and histologic response. We then evaluated the relationships between DKK1 expression in biopsy samples with patient outcomes in two public OS cohorts. Finally, we integrated single-cell transcriptomic analyses with immunohistochemical staining of patient tumour specimens to characterize the cellular distribution of DKK1 and identify candidate cell populations involved in its signalling. Together, these analyses aimed to characterize the dynamics of circulating DKK1, evaluate the prognostic relevance of tumour DKK1 expression, and investigate its cellular distribution and potential signalling context within the OS microenvironment.

## Results

### Plasma DKK1 declines steadily during treatment

The study cohort comprised 34 patients with OS who were enrolled in a clinical trial, MCC20320, designed to identify and evaluate blood-based biomarkers of sarcoma prognosis and response to therapy. The mean enrolment age of the OS cohort was 15.5 years. Metastatic status at diagnosis was documented for 33 patients, while histologic response was available for 27; seventeen healthy donors provided comparing samples (Table 1). In patients, blood was collected at defined stages of therapy, during post-therapy follow-up, and at the time of relapse, processed to isolate plasma, and DKK1 concentration was measured using a Luminex immunoassay (Fig. 1A).

**Figure 1:**
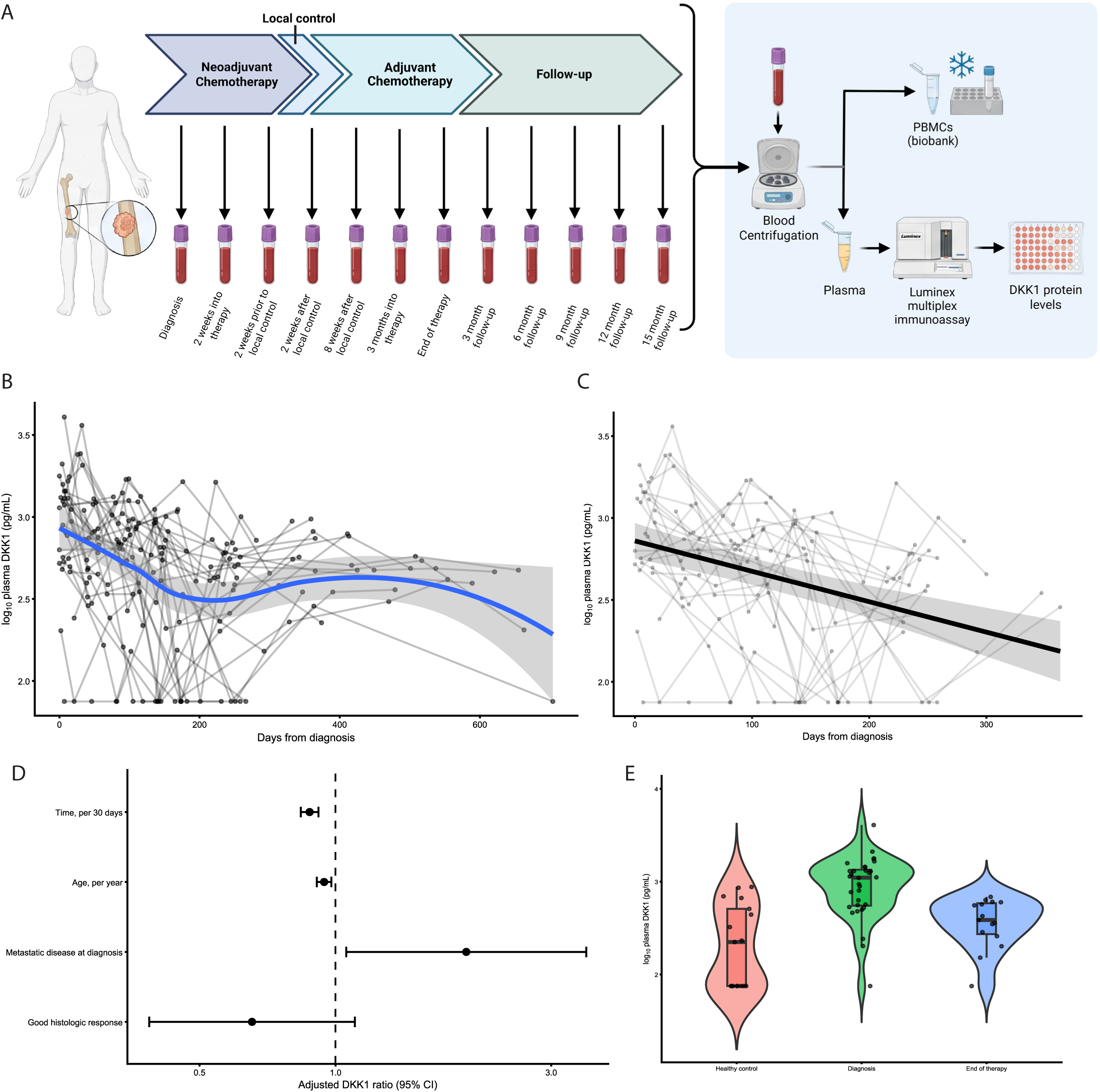
Plasma DKK1 declines during therapy. **A)** Schematic of study design. Created in BioRender. Uceda Arias-Stella, E. (2026) https://BioRender.com/f8xb2b9 **B)** Individual longitudinal plasma DKK1 measurements plotted against days from diagnosis. Lines connect measurements from the same patient; the blue line and shaded region show the LOESS fit and 95% confidence interval, respectively. **C)** Individual trajectories from diagnosis through end of therapy with the adjusted population-level prediction from the linear mixed-effects model (LMM) and 95% confidence interval. **D)** Adjusted DKK1 ratios and 95% confidence intervals for fixed effects in the diagnosis-to-end-of-therapy LMM. The effect of time is expressed per 30 days; the dashed vertical line indicates a ratio of 1. **E)** Distribution of log10-transformed plasma DKK1 measurements in healthy controls and in patients with OS at diagnosis and end of therapy.

**Table 1:** Clinical cohort characteristics.

| Characteristic | OS cohort (N = 34) | Healthy controls (N = 17) |
|---|---|---|
| Age at enrollment, mean ± SD | 15.5 y ± 5.7 | 26 ± 3.9 |
| Sex |  |  |
| Male, n (%) | 18 (53) | 10 (59) |
| Female, n (%) | 16 (47) | 7 (41) |
| Metastasis at diagnosis |  |  |
| Yes, n (%) | 4 (12) | - |
| No, n (%) | 29 (85) | - |
| *NA*, n (%) | 1 (3) | - |
| Histologic response |  |  |
| % Necrosis ≥90, n (%) | 11 (32) | - |
| % Necrosis <90, n (%) | 16 (47) | - |
| *NA*, n (%) | 7 (21) | - |

Individual plasma DKK1 measurements varied widely across patients and over time, showing an initial decline followed by a plateau and potential rebound after end of therapy (Fig. 1B). A generalized additive mixed model (GAMM) fit confirmed the nonlinearity of the trajectory (Supp. Fig. 1C); however, fewer samples were collected after end of therapy, which increased the uncertainty in the trajectory of the latter part of the curve. Therefore, we examined the interval between diagnosis and end-of-therapy separately, during which the plasma DKK1 declined approximately linearly (Fig. 1C).

We modelled the diagnosis-to-end-of-therapy interval using a multivariable linear mixed model (LMM) adjusted for age, metastatic status at diagnosis, and histologic response (138 observations from 26 patients, Fig. 1D). In the primary restricted maximum likelihood (REML) fit, DKK1 declined at a rate of -0.00192 log10 units per day, or around 12.4% per 30-day period. Older age was associated with lower DKK1, at a rate of 5.7% per year (Table 2). The estimated time effect remained negative under alternative approaches to handling measurements below the assay’s detection limit (Supp. Fig. 1F).

**Table 2:** Linear mixed model fit for samples collected between diagnosis and end of therapy.

| **Fixed effect** | **REML estimate<br>(95% CI)** | **P** | **ML estimate<br>(95% CI)** | **P** |
|---|---|---|---|---|
| Intercept | 2.93<br>(2.67 – 3.19) | 1.81×10<sup>-15</sup> | 2.92<br>(2.718 – 3.126) | 2.00×10<sup>-16</sup> |
| Days from diagnosis | -0.00192<br>(-0.00257 – -0.00126) | 6.20×10<sup>-8</sup> | -0.00186<br>(-0.00250 – -0.00121) | 8.03×10<sup>-8</sup> |
| Age, centered at median | -0.0253<br>(-0.0426 – -0.00808) | 0.00635 | -0.0263<br>(-0.0396 – -0.0129) | 3.50×10<sup>-4</sup> |
| Metastatic at diagnosis | 0.289<br>(-0.00184 – 0.580) | 0.0513 | 0.278<br>(0.0627 – 0.494) | 0.0139 |
| Histologic response: ≥90% necrosis | -0.185<br>(-0.428 – 0.0585) | 0.128 | -0.201<br>(-0.390 – -0.0127) | 0.0372 |
| **Random effect** | **REML variance** | **SD** | **ML variance** | **SD** |
| Patient intercept | 0.0219 | 0.148 | 0.00609 | 0.0781 |
| Residual | 0.102 | 0.320 | 0.101 | 0.318 |
| **Overall model fit** | **REML** |  | **ML** |  |
| Marginal *R<sup>2</sup>* | 0.380 |  | 0.408 |  |
| Conditional *R<sup>2</sup>* | 0.489 |  | 0.442 |  |
| Number of patients | 26 |  |  |  |
| Number of observations | 138 |  |  |  |

DKK1 levels at diagnosis and end-of-therapy were higher than those observed in healthy controls (Fig. 1E). These comparisons were interpreted descriptively and with caution, as healthy donors were older than the patient cohort, and patient and control samples were collected and processed differently and assayed on different plates, with internal and quality controls varying between plates, with coefficients of variation of 28-89% (Table 1; Supp. Fig. 1D and 1E). The restricted longitudinal analysis from diagnosis to end-of-therapy therefore provides the main evidence for changes in plasma DKK1 during treatment.

### Local control does not result in a detectable decrease in DKK1 levels

To assess whether surgical resection of the tumour was associated with changes in circulating DKK1 levels, we aligned serial measurements to local control. DKK1 concentrations decreased across this interval without a clear abrupt shift in the trajectory (Fig. 2A). Paired comparisons detected no difference between the samples obtained pre- and post-surgical resection of the primary tumour, with the samples obtained prior to local control and two (p = 0.93, 24 pairs) and eight weeks (p = 0.21, 22 pairs) afterward (Fig. 2B).

**Figure 2:**
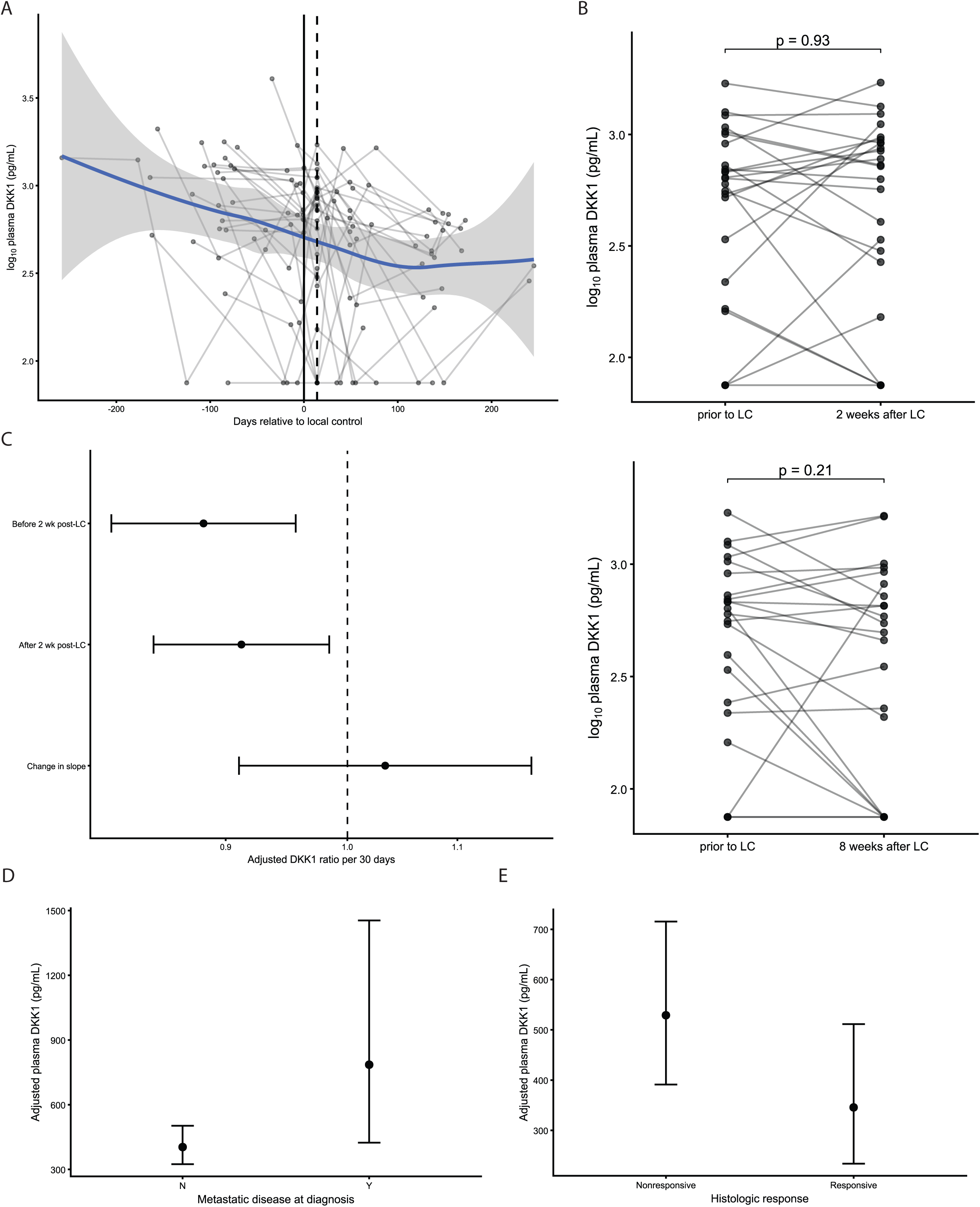
Circulating DKK1 shows no detectable change in trajectory around local control. **A)** Individual plasma DKK1 trajectories aligned to the estimated date of local control (day 0) for samples collected between diagnosis and end of therapy. The vertical lines denote local control and the first scheduled post-local-control collection at 14 days. The blue line and shaded region show a LOESS fit and 95% confidence interval. **B)** Paired plasma DKK1 measurements obtained before local control and at 2 weeks (top) or 8 weeks (bottom) after local control. Lines connect measurements from the same patient; brackets indicate paired t-test comparisons. **C)** Adjusted DKK1 ratios per 30 days for the two segments of the piecewise mixed-effects model and the corresponding slope-change term, with 95% confidence intervals. The dashed vertical line indicates a ratio of 1. **D)** Back-transformed model-adjusted plasma DKK1 estimates according to metastatic disease at diagnosis. Points and error bars represent adjusted estimates and 95% confidence intervals from the diagnosis-to-end-of-therapy LMM. **E)** Back-transformed model-adjusted plasma DKK1 estimates according to histologic response. Points and error bars represent adjusted estimates and 95% confidence intervals from the diagnosis-to-end-of-therapy LMM. Histologic response was defined as ≥90% tumor necrosis.

We also fitted a piecewise LMM, with a knot at the first scheduled post-surgery sample, 14 days after the estimated date of surgery (defined as 14 days before the two week post-surgical sample was obtained). In this analysis, both the earlier and later segments show a decline in DKK1 levels, with no significant difference in their slopes (Fig. 2C); analyses using alternative windows around local control likewise did not identify a clear change in slope (Supp. Fig. 2D). Together, these analyses did not identify a clear change in circulating DKK1 around local control at the sampled timepoints.

### Metastatic status at diagnosis and poor histologic response are associated with higher estimated circulating DKK1 levels

We next examined the relationship between clinical features and DKK1 concentrations during treatment. Patients with metastatic disease at diagnosis had a higher estimated DKK1 concentration than those with localized disease (ratio 1.95, 95% CI 0.996-3.80, p = 0.051; Fig. 2D). Given the small number of patients presenting with metastatic disease in the primary model (n = 3), we performed a sensitivity analysis using an expanded cohort that included patients with incomplete percent necrosis data (162 observations from 32 patients, 4 with metastatic disease at diagnosis). The expanded cohort model similarly showed increased DKK1 concentrations in patients with metastatic disease (beta: 0.305, ratio: 2.02, 95% CI 1.10-3.71, p = 0.026). In addition, leave-one-out analyses in both the primary and expanded-cohort models showed consistent directionality in the relationship between metastatic status at diagnosis and DKK1 levels, indicating that the estimated direction of this association was not driven by any single patient (Supp. Fig. 2E).

Neither diagnostic DKK1 concentrations nor their change from diagnosis to pre-surgical resection show a clear relationship with percent tumour necrosis (treated as a continuous variable) in exploratory analyses (Supp. Fig. 2F). Comparisons using a 90% necrosis threshold similarly did not identify differences in diagnostic DKK1 concentrations (p = 0.73); however, there appeared to be a difference in preoperative change in DKK1, though this only approached significance (p = 0.10; Supp. Fig. 2G). In the primary model, patients with at least 90% tumour necrosis (deemed as good responders to chemotherapy) had lower estimated DKK1 concentrations compared to patients with less than 90% necrosis (deemed as poor responders), though this association was also more uncertain (ratio 0.653, 95% CI 0.373-1.14, p = 0.128; Fig. 2E). Nevertheless, leave-one-out analyses of the primary model showed consistent directionality in the relationship between histologic response and DKK1 levels (Supp. Fig. 2H). Overall, this suggests that patients with good histologic response have lower adjusted DKK1 concentrations. Although the confidence interval included no difference, the consistency of this estimate across leave-one-out analyses supports further investigation.

### Higher tumour DKK1 expression is associated with poor outcomes

To examine the clinical relevance of DKK1 expression in primary tumours, we analysed diagnostic biopsy tumour expression data and outcomes from TARGET-OS and GSE33382 (Table 3) [19, 20]. *DKK1* expression was standardised within each cohort and modelled using Cox regression; cohort-specific estimates were combined by inverse-variance fixed effect pooling (Fig. 3A). For overall survival, the association estimates were similar in both cohorts, with the pooled analysis showing higher DKK1 expression is associated with a 35% increased hazard of death (95% CI, 2-78%, p = 0.034; Fig. 3B). Similarly, relapse-free/metastasis-free survival (RMFS) analyses showed that higher *DKK1* expression was associated with increased hazard of metastasis in the TARGET-OS cohort (HR, 1.99, 95% CI 1.16-3.42, p = 0.0120), though the estimate in GSE33382 was smaller and less precise (HR, 1.22, 95% CI 0.79-1.88, p = 0.361). The pooled estimate suggested that higher *DKK1* expression is associated with a 48% increased hazard of metastasis (95% CI, 6-107%, p = 0.023; Fig. 3C).

**Figure 3:**
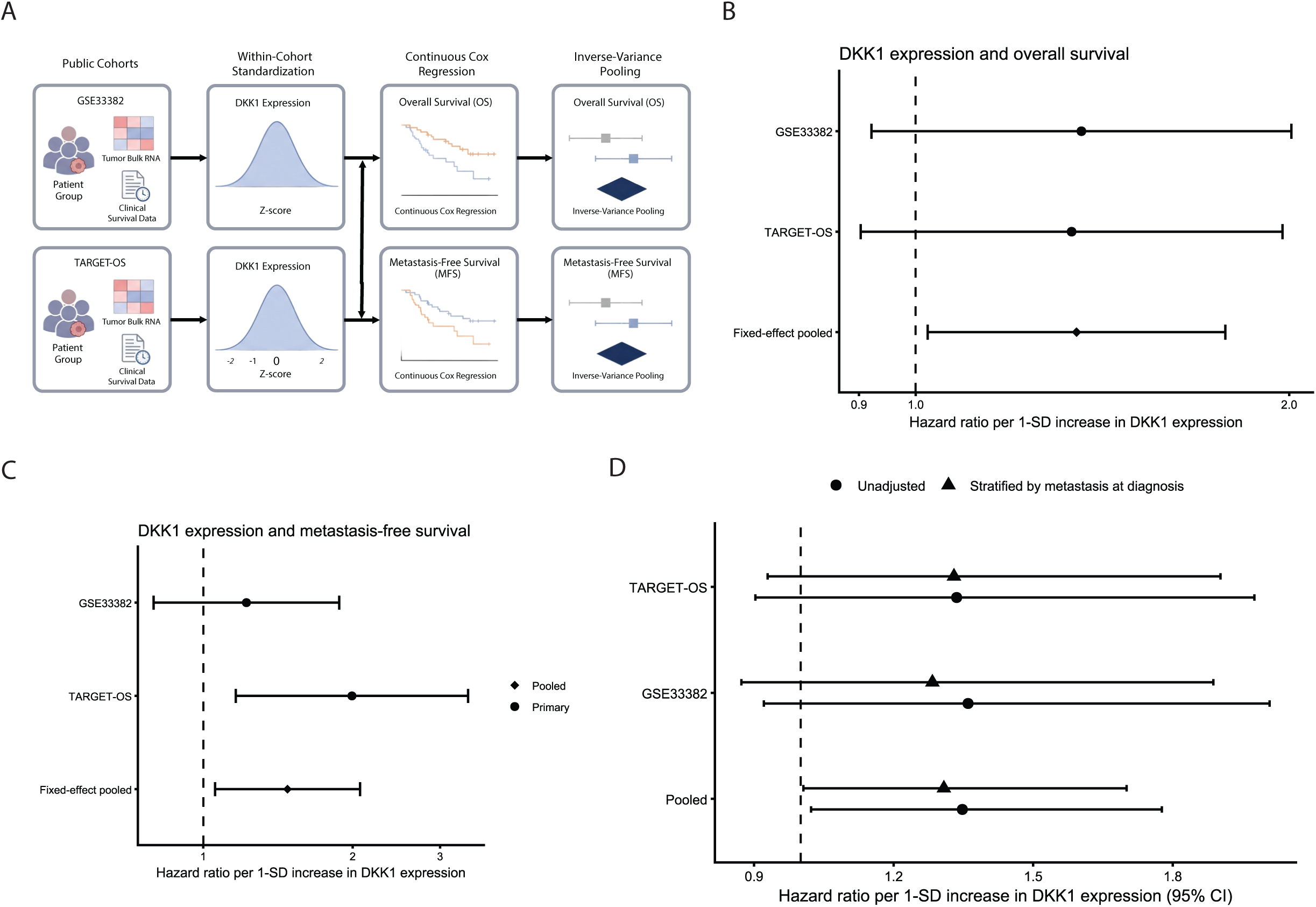
Tumor DKK1 expression is associated with adverse outcomes in independent OS cohorts. **A)** Schematic of public cohort survival analysis workflow. Created with BioRender. Uceda Arias-Stella, E. (2026) https://BioRender.com/ze03yuh **B)** Cohort-specific and pooled overall survival hazard ratios per 1-SD increase in DKK1 expression, with 95% confidence intervals. Pooled estimates were calculated using fixed-effect inverse-variance weighting. **C)** Cohort-specific and pooled metastasis-free survival HRs per 1-SD increase in DKK1 expression, with 95% confidence intervals; MFS analysis was restricted to patients with localized disease at diagnosis. Pooled estimates were calculated using fixed-effect inverse-variance weighting. **D)** Comparison of unadjusted OS estimates with estimates from Cox models stratified by metastatic status at diagnosis. Pooled estimates were calculated using fixed-effect inverse-variance weighting.

**Table 3:** Clinical characteristics of GSE33382 and TARGET-OS cohorts.

| **Characteristic** | **GSE33382 (N = 83)** | **TARGET-OS (N= 87)** |
|---|---|---|
| Age at diagnosis, median (IQR) | 16.0 (13.7–18.8) | 14.5 (12.3–17.3) |
| Male, n (%) | 55 (66.3) | 50 (57.5) |
| Female, n (%) | 28 (33.7) | 37 (42.5) |
| Metastatic at diagnosis, n (%) | 14 (16.9) | 22 (25.3) |
| Localized at diagnosis, n | 69 | 65 |
| Deaths, n | 25 | 29 |
| Median OS, months (95% CI) | NR | 126.9 (94.7–NR) |
| MFS evaluable, n | 68 | 63 |
| Metastatic events after diagnosis, n | 20 | 13 |
| Median MFS, months (95% CI) | NR | NR |

Survival medians estimated by Kaplan–Meier analysis. NR, not reached. MFS restricted to patients with localized disease at diagnosis.

When the overall survival analyses allowed separate baseline hazards according to metastatic status at diagnosis, the *DKK1* effect estimates remained above one, with wide confidence intervals in the individual cohorts (Fig. 3D). These results support an association between higher tumour *DKK1* expression and adverse outcomes. Kaplan–Meier visualizations and model diagnostics are provided in Supp. Fig. 3.

### DKK1 in osteosarcoma is expressed predominantly by malignant cells

We next decided to investigate the cellular origins of DKK1 expression in primary OS. We leveraged the OsteoCAR atlas, a cross-species collection of scRNA-seq datasets that have been integrated and annotated [21]. We selected primary-site tumours, including local recurrences, from two human datasets (BC and OS). *DKK1* expression was highly heterogeneous across different tumours (Supp. Fig. 4A), but the majority of detected *DKK1* transcripts originated from malignant cells (Fig. 4A; Supp Fig. 4A–B). Two exceptions were BC21 and OS5, in which nonmalignant cells accounted for a greater proportion of *DKK1* transcripts. However, these were the lowest DKK1-expressing tumours in their respective datasets, and contained fewer than 10 raw DKK1 pseudobulk counts, limiting interpretation.

**Figure 4:**
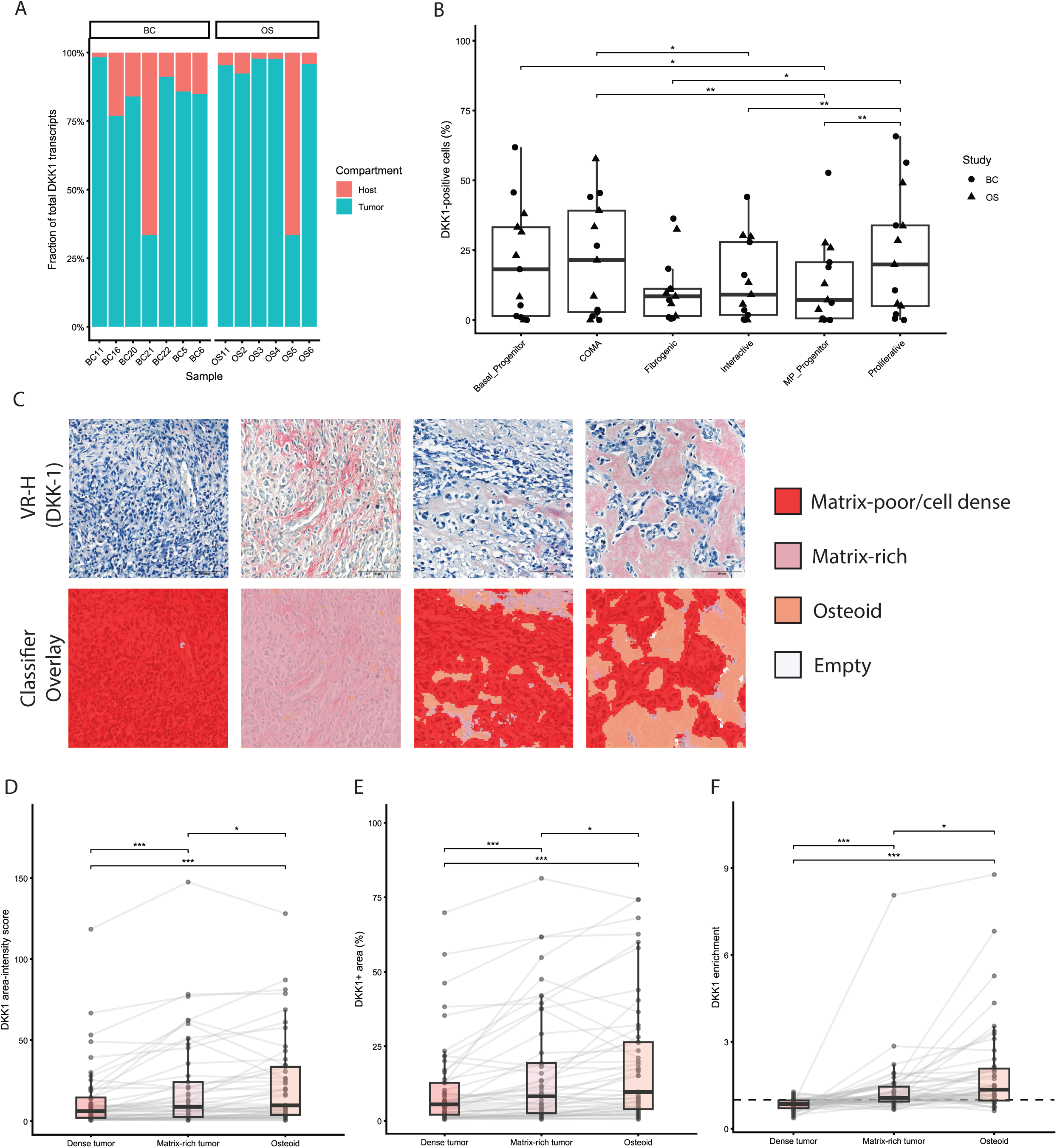
Tumoral DKK1 is localized to osteoid- and matrix-rich regions. **A)** Fraction of DKK1 transcripts expressed in either nonmalignant/host or tumoral compartments by sample from two independent scRNA-seq studies focusing on primary tumours. **B)** Proportion of DKK1+ tumor cells by OsteoCAR Level 2 annotation across both studies. **C)** Representative images from OS tissue microarray slide stained with anti-DKK1 antibody and hematoxylin (top), and QuPath classifier overlay showing assignment to three classifier compartments: matrix-poor/cell dense, matrix-rich, and osteoid (bottom). **D)** DKK1 area intensity score across the three QuPath classifier compartments. Lines connect compartments from the same TMA core. **E)** DKK1+ area fraction per TMA core across the three QuPath classifier compartments. Lines connect compartments from the same TMA core. **F)** DKK1 enrichment score per TMA core across the three QuPath classifier compartments. Lines connect compartments from the same TMA core.

Within the malignant compartment, the fraction of *DKK1*-expressing cells was significantly different across OsteoCAR-defined subpopulations (p = 0.0003; Fig. 4B). Basal progenitor, proliferative, and COMA (chondroid-osteoid matrix-associated) subpopulations had higher proportions of *DKK1*-positive cells compared to the fibrogenic, interactive, and MP progenitor subpopulations. Notably, the OsteoCAR atlas describes the basal progenitor and proliferative subpopulations as enriched in regulons that suggest a less differentiated phenotype, while the COMA subpopulation is described as having a more differentiated phenotype and being enriched in hypoxia- and angiogenesis-related genes, as well as having a spatial association with chondroid-osteoid matrix deposition [21]. Overall, this suggests that *DKK1* is expressed mainly by malignant cells, but that expression is distributed across multiple malignant cell states.

### DKK1 protein is enriched in osteoid and matrix-rich regions of primary tumours

Given the heterogeneous distribution of *DKK1* transcript across malignant cell states, and the possibility that secreted DKK1 protein may accumulate at sites far from its cellular site of production, we decided to explore protein expression in primary tumour samples. We performed immunohistochemical staining of DKK1 on a tissue microarray (TMA) containing 50 cores from primary OS tumours. Using QuPath, we developed a pathologist-guided pixel classifier to distinguish three histologic tumour compartments: matrix-poor/cell dense regions, matrix-rich tumour regions, and osteoid (Fig. 4C). Consistent with these compartment assignments, cell dense regions exhibited higher nuclear density compared to matrix-rich and osteoid regions (Supp. Fig. 4C).

Of the 50 TMA cores, 49 contained all three histologic compartments and were included in the paired compartment-level comparisons. DKK1 staining differed significantly across histologic compartments (Friedman test, p = 4.07 × 10⁻⁷; Fig. 4D). The DKK1 area-intensity score was highest in osteoid, followed by matrix-rich tumour and cell-dense regions, with post hoc Conover comparisons identifying significant differences between compartments. A similar pattern was observed for the fraction of DKK1-positive area within each compartment (Friedman test, p = 5.20 × 10^⁻7^; Fig. 4E). Because the relative areas occupied by each histologic compartment varied between TMA cores (Supp. Fig. 4D), we also calculated DKK1 enrichment as the fraction of total DKK1-positive area attributable to a compartment divided by that compartment’s fraction of total tissue area. DKK1 enrichment also differed significantly between compartments (p = 5.20 × 10^⁻7^), with the greatest enrichment in osteoid, followed by matrix-rich and cell-dense regions (Fig. 4F). Taken together, these findings suggest that, although *DKK1* transcripts are detected predominantly in malignant cells, DKK1 protein expression is preferentially distributed within osteoid and matrix-rich regions of primary tumours.

### No significant increase in plasma DKK1 was detected at relapse

Given the association between tumoral *DKK1* expression and metastatic hazard, we next investigated whether circulating DKK1 changed upon disease relapse. Plasma DKK1 concentrations were higher at relapse compared with end-of-therapy samples, although this was not statistically significant (Fig. 5A). As relapse occurred at different timepoints during treatment or follow-up across patients, we compared each patient’s relapse sample with their most recent available pre-relapse sample (Fig. 5B). This paired analysis revealed no significant change in circulating DKK1 at relapse, consistent with our finding that circulating DKK1 does not behave like a real-time readout of tumour burden.

**Figure 5:**
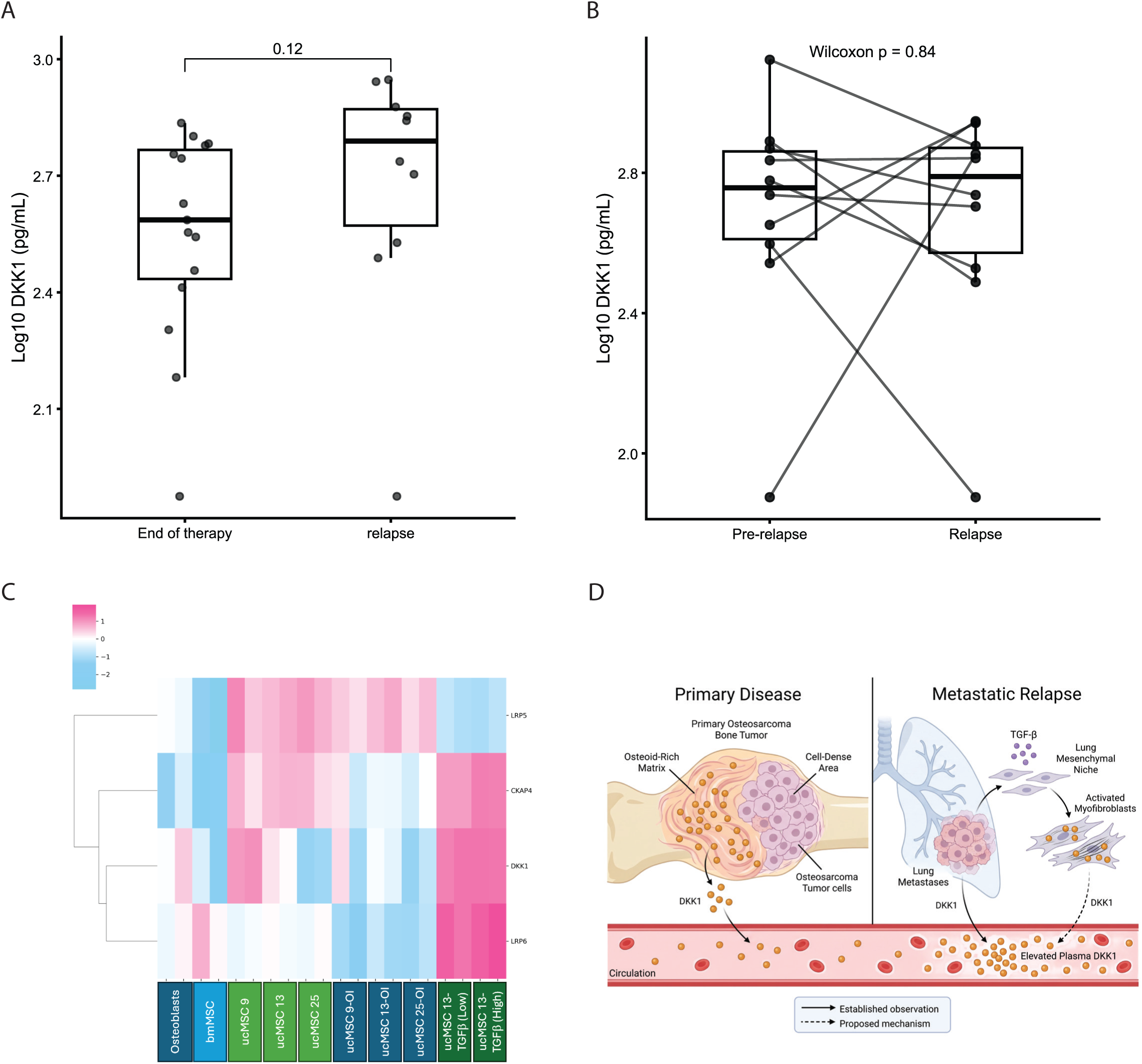
Circulating DKK1 at relapse and potential contributions from nonmalignant mesenchymal cells. **A)** Distribution of plasma DKK1 concentrations in samples collected at the end of therapy and at disease relapse. **B)** Paired comparison of plasma DKK1 concentrations at relapse and the most recent available pre-relapse sample from the same patient. Lines connect paired measurements. **C)** Heatmap showing relative transcript expression of DKK1 and selected DKK1-associated receptors (LRP5, LRP6, and CKAP4) in human osteoblasts, mesenchymal stem cells, and umbilical cord-derived mesenchymal stem cells (ucMSCs) under basal conditions, following osteogenic differentiation, or cultured in conditioned medium from control or TGF-β-overexpressing LNCaP cells. **D)** Working model integrating the observed distribution of DKK1 transcripts and protein with proposed contributions from malignant and nonmalignant compartments. Contributions of these compartments to circulating DKK1, including those of mesenchymal cells within pulmonary metastatic niches, remain to be established. Created in BioRender. Uceda Arias-Stella, E. (2026) https://BioRender.com/gs84x27

### DKK1 expression in cultured mesenchymal cells identifies a candidate nonmalignant source

Finally, given the potential contribution of nonmalignant cells to circulating DKK1, particularly at the time of relapse, we explored whether *DKK1* expression could be associated with a reactive stromal phenotype. We analysed transcriptomic data from primary osteoblasts and human umbilical cord-derived mesenchymal stem cells (ucMSCs) cultured under basal conditions, induced towards osteogenic differentiation, or cultured in conditioned medium from control or TGF-β-overexpressing LNCaP cells to promote a reactive, myofibroblast-like phenotype. *DKK1* expression was elevated in the TGF-β-exposed ucMSCs compared with other conditions (Fig. 5C). This suggests that *DKK1* expression may be associated with TGF-β-induced MSC activation, identifying a potential nonmalignant source that could augment the DKK1 levels that derive from the malignant tumour cells.

## Discussion

Circulating DKK1 declines over treatment course, with no noticeable abrupt change around the time of surgical resection of the primary tumour. Previous studies have shown elevated DKK1 serum levels at diagnosis in OS patients, as well as protein expression in the tumour [15]. These findings extend earlier cross-sectional observations of elevated serum DKK1 in OS by characterizing its trajectory during treatment. We also show that higher diagnostic tumour *DKK1* expression is associated with poorer survival outcomes and with increased metastatic risk. Our analysis of single-cell transcriptomic datasets show that *DKK1* transcripts are predominantly attributable to malignant cells within primary tumours, with the protein being enriched in matrix-rich and osteoid compartments of the tumour. Taken together, these observations suggest a unique role of DKK1 at the interface between malignant-cell biology and the surrounding bone and stromal environment, although the separate study cohorts prevent direct assessment of how tumour expression and tissue localization relate to circulating concentrations.

The main finding that DKK1 levels decline monotonically during treatment raises the possibility of multiple potential DKK1 sources. Chemotherapy-induced cell death likely plays a role in this reduction, as the decline in DKK1 precedes surgical resection of the tumour, while host responses to the tumour, treatment, and tissue repair may influence subsequent concentrations. In addition, the resection itself does not seemingly alter the course of the decline, which is compatible with an additional, nonmalignant source of circulating DKK1.

Recent work in bone-metastatic breast cancer models has shown that bone-derived DKK1 is the predominant source of systemic DKK1, while cancer-associated fibroblasts (CAFs) contribute most of the tumoral DKK1 in the primary lesion [17]. In OS, reactive changes in the surrounding bone and postoperative repair are therefore plausible contributors. Platelets are another likely nonmalignant source, as it has been shown that DKK1 protein is present in platelet extracts [16, 22]. Interestingly, *DKK1* mRNA is not expressed in platelets, suggesting that DKK1 protein presence in platelets is due to uptake by endocytosis [22]. Thus, platelets may provide a releasable pool of DKK1 without identifying the tissue in which the protein was originally synthesized.

We also explored the relationship between tumour *DKK1* expression and patient survival, and show that a continuous expression analysis supports a negative association between *DKK1* and both overall survival and relapse-free/metastasis-free survival. Notably, there is evidence that DKK1-targeted therapy reduces ALDH activity, enhances osteogenic differentiation in the primary tumour, reduces metastatic burden in OS models [11-13, 23]. Wnt inhibition through DKK1 expression has also been shown to promote a quiescent, immune-evading state in lung and breast cancer models, suggesting that DKK1 can influence persistence and immune escape, which are critical for the establishment of a metastatic niche [24].

An additional finding of this study was the potential relationship between circulating DKK1 and histologic response to neoadjuvant chemotherapy. DKK1 concentrations at diagnosis were not significantly associated with subsequent tumour necrosis, suggesting that baseline DKK1 alone is not a predictor of histologic response. However, when longitudinal measurements were considered, patients with good histologic response had lower adjusted DKK1 concentrations throughout treatment, albeit with some uncertainty. These findings raise the possibility that changes in circulating DKK1 during neoadjuvant chemotherapy may be more informative about treatment-associated biological responses than its concentration at diagnosis. Experimental work linking DKK1 to ALDH-associated stress-tolerance provides a biological rationale for examining the relationship of DKK1 to chemoresistance [23]. Larger prospective cohorts with serial sampling and detailed histologic response data will be required to determine whether early changes in DKK1 provide information about chemotherapy response beyond baseline measurements.

*DKK1* expression across several malignant cell states suggests that it is not restricted to a single differentiation programme. This finding fits with recent studies showing that OS comprises heterogeneous mesenchymal and osteoblast-associated states that can coexist within individual tumours, with subpopulations existing along multiple potential mesenchymal lineages [25, 26]. Our analysis of scRNA-seq datasets show both progenitor-like tumour cells and more differentiated matrix-associated tumour cells express *DKK1*. Earlier work has also shown that *DKK1* is regulated by the osteoblast transcription factor Sp7, while DKK1 immunodepletion attenuates the inhibition of osteogenesis in MSCs cultured in OS-conditioned medium [9, 15]. Osteoid production, thus, need not imply completion of terminal osteogenic differentiation, and DKK1 may participate in paracrine regulation within a heterogeneous tumour. These possibilities require functional testing, particularly as the consequences of Wnt signalling repression differ across OS models [27, 28].

Our IHC analysis shows a clear enrichment of DKK1 protein along matrix-rich and osteoid regions of the tumour, adding a spatial dimension to this interpretation. Importantly, protein localization need not coincide with the site of transcription, as secretion, extracellular association, and turnover can influence local availability. This could point to the osteoid matrix being a DKK1 reservoir, and DKK1-glycosaminoglycan interactions have been identified, showing this extracellular association is plausible [29]. The discordance between cellular transcript expression and extracellular protein distribution also raises the possibility that OS cells produce DKK1 to modulate differentiation within the tumour environment. However, our staining does not establish a functional reservoir or identify the cells that produce or respond to the protein. Combined spatial transcriptomics and histological DKK1 staining could be used to assess these hypotheses, and would allow us to deconvolute candidate DKK1 sources and protein localization. In addition, spatial transcriptomic analysis could reveal receptor localization and downstream pathway readouts, allowing us to distinguish production from accumulation and identify candidate paracrine relationships. Recent spatial transcriptomic studies in OS demonstrate the feasibility of this approach [30].

Our observation of increased DKK1 expression in MSCs treated with cultured media from TGF-β-overexpressing LNCaP cells provides another link between potential host sources of DKK1 and tumour-associated tissue responses. TGF-β signalling is a mediator of wound response stromal remodelling and has been shown to induce a fibrotic response in mesenchymal stem cells, including lung-resident MSCs [31-33]. Within this framework, the response of MSCs to TGF-β stimulation in the context of the profibrotic microenvironment could be associated with pulmonary metastatic colonization. Notably, recent work has shown that disseminated OS cells induce a persistent wound-healing response in the lung, accompanied by profibrotic epithelial and macrophage states as well as extensive ECM deposition [34]. Within this framework, our results raise the possibility that mesenchymal cells responding to tissue injury or profibrotic signals in the lung might provide an additional source of DKK1. This possibility informs our working model (Fig. 5D), in which DKK1 production and local availability reflect contributions from tumour cells and responsive host compartments. Such contributions could help explain why circulating concentrations do not change directly with removal of the primary tumour. However, whether analogous DKK1-producing mesenchymal states occur within pulmonary metastatic niches and whether their DKK1 contributes to circulating levels remains to be established.

Our findings have different implications for biomarker development and prognostics. The association between *DKK1* expression and adverse outcomes supports further evaluation of *DKK1* as a prognostic marker. In contrast, plasma DKK1 does not behave like a real-time readout of tumour burden, so its utility for monitoring tumour burden or chemotherapy response, or for detecting disease progression, remains uncertain. Prospective studies should evaluate whether early changes in plasma DKK1 improve assessment of treatment response. The associations presented here also complement existing preclinical evidence that DKK1-targeted interventions can reduce OS progression and metastasis [11-13]. Receptor context may be important to this approach, because DKK1 can activate CKAP4-dependent PI3K signalling as well as antagonize canonical Wnt signalling [35]. These findings provide a rationale for further therapeutic investigation; receptor expression and downstream pathway activity may clarify the biological consequences of DKK1 inhibition.

Several limitations constrain the interpretation of the plasma DKK1 findings. Although repeated sampling allowed assessment of within-patient changes, the total number of patients was modest, particularly those with metastatic disease at diagnosis. In addition, missing histologic-response data reduced the multivariable analysis cohort. Associations with metastatic disease and tumour necrosis thus require confirmation with larger sample sizes. Surgery dates were not available for analysis, so they were estimated using the post-surgical sample draw dates, so it is possible that immediate abrupt changes in DKK1 occurring post-operatively were missed.

Furthermore, the plasma, bulk transcriptomic, single-cell transcriptomic, and histological analyses involved separate cohorts, preventing direct assessment of how tumoral DKK1 expression and protein localization relate to circulating concentrations in individual patients. Bulk expression may reflect cellular composition as well as expression within individual cells, while single-cell estimates depend on cell recovery, transcript detection, and annotation. This is particularly important for DKK1, whose expression may be low and transient. Likewise, expression in cultured mesenchymal cells demonstrates a potential source under the tested conditions but does not quantify its contribution in patients. Paired longitudinal sampling, spatial RNA–protein analysis, and functional perturbation will be needed to test the proposed relationships among DKK1 production, extracellular localization, and tumour–host signalling.

## Methods

### Study design

Patients in the plasma cohort were enrolled in an ongoing multi-centre clinical trial, MCC20320, a study evaluating blood-based biomarkers of disease response and prognosis in children, adolescents, and young adults with sarcomas. This multi-centre clinical trial was approved by the Advarra central Institutional Review Board, and was conducted in accordance with the Ethical Principles and Guidelines for the Protection of Human Subjects of Research. Newly diagnosed patients less than or equal to 40 years old and weighing over 8 kg were enrolled at participating centres. Plasma from clinical waste of presumed healthy individuals was used for healthy controls. Study accrual opened in 2019 and is ongoing. This report focuses on subjects belonging to Cohort 4 of the trial, which is defined as: confirmed patients with osteosarcoma.

### Plasma collection and processing

Blood was collected at enrolment in a Streck Cell-free DNA BCT® tube and shipped to the University of Colorado Sarcoma Research Lab in ATS1 temperature-regulated containers (Akuratemp, Arden, NC, USA). Plasma was isolated using double-centrifugation and was frozen at −80 °C until use, as previously described [36]. Samples were aliquoted and frozen at −80 °C and underwent no more than two freeze/thaw cycles.

### Luminex assays

All samples were measured in technical duplicates and averaged for a final concentration. DKK1 protein was measured using HDKK1-MAG obtained from Sigma Millipore (St. Louis, MO, USA). Kit was used according to the manufacturer’s instructions. Data were acquired on a Luminex Magpix XMAP Multiplex Reader (Luminex Technologies (Madison, WI, USA), RRID:SCR_023348) and were analyzed using Belysa Immunoassay Curve Fitting Software (Millipore Sigma, Burlington, MA, USA). Interpolated concentrations of analyte were log-transformed before analysis to reduce non-normal skew and inter-assay variability.

### Public transcriptomic dataset mining and survival analyses

Publicly available osteosarcoma transcriptomic and clinical data were obtained from GSE33382 and the TARGET-OS cohort. *DKK1* expression was standardized within each dataset to a mean of zero and standard deviation of one. Associations between *DKK1* expression and overall survival (OS) or metastasis-free survival (MFS) were assessed using Cox proportional hazards regression. MFS analyses were restricted to patients without metastatic disease at diagnosis. Hazard ratios (HRs) were reported per standard-deviation increase in *DKK1* expression. Cohort-specific estimates were combined using fixed-effect inverse-variance meta-analysis. Kaplan– Meier curves were generated using cohort-specific median DKK1 expression to define high- and low-expression groups.

### scRNA-seq analysis

Publicly available single-cell RNA sequencing (scRNA-seq) data were obtained from the OsteoCAR atlas [21], using the annotated Seurat objects deposited in Figshare (doi:10.6084/m9.figshare.31029559, accessed May 15, 2026). Human osteosarcoma specimens were selected from the atlas’s primary-site dataset and restricted to two independent studies: BC (GSE152048) and OS (GSE162454). The analysis included 7 specimens from BC and 6 from OS, comprising 85224 and 44153 cells respectively. Samples from other studies and metastatic sites were excluded. Cell classifications were obtained from the original OsteoCAR annotations, using Level 0 to distinguish malignant tumour cells from nonmalignant host cells and Level 2 to identify specific cell populations and malignant transcriptional states.

Raw transcript counts were extracted from the Seurat RNA assay. DKK1 counts were summed across cells belonging to each Level 2 population to generate per-sample cell-state-level pseudobulk counts. The relative contribution of each population was calculated by dividing its summed DKK1 counts by the total DKK1 counts detected across all malignant and nonmalignant cells from the same specimen. Specimens with no detected DKK1 transcripts were assigned undefined rather than zero source fractions. For comparisons of DKK1 expression between specimens, raw counts from malignant cells were summed by gene within each specimen to generate sample-level pseudobulk profiles. DKK1 expression was normalized to counts per million (CPM), using the total number of transcripts detected across all genes in the malignant compartment as the denominator, and transformed as log₂(CPM + 1).

The proportion of DKK1-expressing cells was assessed using log-normalized RNA expression data. Cells with detectable DKK1 expression (DKK1 > 0) were classified as DKK1-positive, corresponding to nonzero raw DKK1 counts. Populations not represented by any recovered cells within a specimen were treated as absent rather than assigned a DKK1-positive fraction of zero. Differences in the percentage of DKK1-positive cells across the six OsteoCAR-defined malignant cell states were assessed using a Friedman test, treating individual specimens as blocks, followed by pairwise Conover comparisons with Holm adjustment for multiple testing.

### Proteolytic in Silico retrieval for DKK1

Optimal proteolytic retrieval was predicted in-silico using the Expasy Peptide Cutter website plug-in on accession number NP_036374.1 as a reference [37]. Given that the primary antibody is polyclonal, trypsin was selected for retrieval since it has the most conservative cleavage profile (29 sites).

### Immunohistochemistry

IHC was performed according to protocols developed for decalcified bone [38]. Paraffin-embedded arrays (TissueArray.com, Osteosarcoma: OS802d, Normal Bone array: BO244g) were pre-warmed in a humidified 100 °C oven for 6 min and de-waxed with xylene. Sections were hydrated through a series of graded ethanol washes: 100% (three times, 3 min each), 95%, 70% (two times, 2 min each). Sections were then washed in GeneTex 1X washing buffer (GeneTex, GTX73344; all wash cycles comprised three washes, 3 min each).

After de-waxing and rehydration, sections were subjected to proteolytic antigen retrieval by incubation with Trypsin (Abcam, ab970) for 7 min at 37°C in a humidified oven. After cooling and washing, endogenous enzymatic alkaline phosphatase (AP) activity was blocked by treating with Bloxall (Vector Laboratories, SP-6000-100) for 10 min at RT. Sections were then washed and incubated in Protein Blocker (GeneTex, GT30963) for 10 min at 37° C, followed by incubation in 5% goat serum (GIBCO, 16-2100-72) diluted in 1X automation buffer (GeneTex, GTX30931), for 30 min at RT. Sections were incubated overnight in primary antibody at 4 °C (Bioss Inc., BS2162R 1:200) diluted in primary antibody diluent (GeneTex, GTX28208). After washing, samples were exposed to biotinylated goat anti-rabbit secondary antibody (Invitrogen 31820, 1:500) diluted in automation buffer for 45 min at 37° C. After washing, sections were then incubated for 30 min in streptavidin-bound AP (Vector Labs, Vectastain ABC kit, AK-5000), followed by washing and 20 min incubation in Vector Red substrate (Vector Labs, SK-5100). Sections were counterstained with Vector hematoxylin QS (Vector Labs, H-3404-100) for 45 seconds at RT, dehydrated through a series of graded ethanol (Two times, two minutes each at 70% and 95%, followed by three times, three minutes at 100%) followed by extensive washing in Histoclear (National Diagnostics, HS-200) and mounting in the non-xylene-based media Omnimount (Electron Microscopy Science, 17997-01).

### TMA image analysis

Images were analyzed using QuPath version 0.6. Stain vectors were estimated empirically from representative regions in the image. Tissue regions were classified according to histologic morphology as dense tumor, matrix-rich tumor, or osteoid. DKK1 immunoreactivity was quantified as the percentage of tissue area exceeding predefined low-, intermediate-, and high-intensity stain thresholds. An area-weighted staining score (range, 0–300) was calculated by summing the three cumulative positive-area percentages. DKK1 enrichment within each compartment was calculated as its proportion of total DKK1-positive area relative to its proportion of total analysed tissue area.

### Cell culture

Umbilical cord-derived Mesenchymal Stem Cells ucMSC9, ucMSC13, ucMSC25 were grown in Alpha MEM + Glutamax media (Gibco, #32561037), containing 10% MSC-grade FBS (Gibco, #12662-029), 0.25X penicillin-streptomycin (Gibco, #15-140-122), and 2 ng/mL Fibroblast Growth Factor (R&D Systems, #233FB025). Cells were passaged at 80% confluency using 0.25% Trypsin (Gibco, #25200-072) at a sub-culture ratio of 1:5.

Human osteoblasts (Promocell) were cultured in Osteoblast Growth medium (Promocell C-27001) supplemented with 0.25X Penicillin-Streptomycin. Human Bone Marrow MSCs (Promocell) were cultured in MSC growth medium 2, according to the manufacturer’s instructions (Promocell C-28009). Both cell lines were sub-cultured around 80% confluency using reagents from the Detach Kit (Promocell C-41200), according to the manufacturer’s instructions. For these cells, RNA was purified from 5 million-cell pellets, using the Qiagen RNeasy mini kit (Qiagen, #74106), according to the manufacturer’s instructions.

LNCaP-PBMN control and LNCaP-TGF-β overexpressing cells were a kind gift from Dr. David Rowley (Baylor College of Medicine, Houston TX). Cells were cultured in RPMI 1640 (Corning, #10040CV) modified to contain 4.5g/L glucose (using Gibco, #A2494001) 2.383 g/L HEPES (Gibco #15630106), and 0.11g/L pyruvate (Gibco #11360070). Basal media was then supplemented with 10% FBS (Corning, #35010CV) and 0.25x Penicillin Streptomycin (Gibco, #15-140-122). Cells were passaged using 0.25% Trypsin (Gibco, #25200-072) at a ratio of 1:5 and allowed for 48hr. Conditioned media were collected from cells around 70% confluency, adding fresh media for a second collection. Conditioned media from both collection time points were pooled and centrifuged to remove cell debris before treatment.

### Induction of osteogenic differentiation

ucMSCs were subcultured into MEMα media (Gibco #12561056) with no nucleosides, in T75 vented flasks and allowed to reach 80% confluency. Media was supplemented with StemXVivo Human Osteogenic Supplement (R&D Systems CCM008), and osteogenesis was induced for 14 days, changing media every two days, according to the manufacturer’s instructions. RNA collection was carried out using the RNeasy Fibrous Tissue Mini Kit (Qiagen 74704), according to the manufacturer’s instructions.

### Induction of myofibroblast phenotype

Cells were seeded in plastic 6-well plates (Fisher, #FB012927) at 50,000 cells per well and allowed to adhere overnight. Cells were treated with 2 mL LNCaP TGF-β (TGF-β-high) or PBMN (TGF-β-low) conditioned media per well for 48 hours. For collection, media was removed from the wells, and 100 microliters of RLT Buffer (Qiagen) was added per well. Cells were scraped into the buffer, and lysates were pooled per condition. RNA was then purified using the Qiagen RNeasy mini kit (Qiagen, #74106), according to the manufacturer’s instructions.

### RNA-seq

RNA-seq library construction and sequencing for two technical replicates was carried out by Genewiz/Azenta (South Plainfield, NJ). Sequence reads were trimmed using Trimmomatic v.0.36. trimmed reads were mapped to the Homo Sapiens GRCh37 reference genome using STAR aligner v.2.5.2b. Unique gene counts were calculated using FeatureCounts from the Subread package v1.5.2. Hit counts were summarized and reported using the gene_id feature in the annotation file. Only reads that fell within exon regions were counted. Heatmaps were generated as previously described [39].

### Statistical analysis

Statistical analyses were performed in R 4.4.3. Longitudinal changes in log₁₀-transformed plasma DKK1 were assessed using linear mixed-effects models, incorporating patient-specific random intercepts to account for repeated measurements. The primary model, for the interval between diagnosis and end of therapy, included days from diagnosis, age, metastatic status at diagnosis, histologic response, and Luminex assay plate as fixed effects. Model coefficients were back-transformed to express adjusted concentration ratios. Concentrations below the assay’s lower limit of detection (150 pg/mL; 26 of 210 patient samples) were set to half the limit (75 pg/mL) before log transformation; alternative substitutions are shown in Supp. Fig. 1F. Models were fitted by restricted maximum likelihood with Satterthwaite degrees of freedom and t-based 95% confidence intervals. All available samples were used in the mixed models, including repeated pre-local-control draws; paired comparisons used the latest sample at each stage.

Nonlinear changes across the complete follow-up period were evaluated using generalized additive mixed models, incorporating a cubic regression spline for time, assay plate, and a patient-specific random effect. Changes around local control were assessed using mixed-effects models with treatment stage as a categorical predictor.

Within-patient changes between diagnosis and subsequent treatment stages were assessed using paired Wilcoxon signed-rank tests, with Hodges–Lehmann estimates and 95% confidence intervals. An exact binomial sign test was additionally used to evaluate the direction of change before local control. Pre-versus post-surgical samples were compared with paired t-tests on log10 concentrations. Relationships between plasma DKK1 and percent tumour necrosis were examined using Spearman rank correlations, and comparisons between histologic response groups used Wilcoxon rank-sum tests.

Differences in DKK1 immunoreactivity across tissue compartments were assessed using Friedman tests followed by pairwise Conover tests. P values were adjusted using the Holm procedure for specified pairwise comparisons and the Benjamini–Hochberg procedure for gene-set analyses. All tests were two-sided, with P < 0.05 considered statistically significant.

## Data Availability

De-identified plasma DKK1 measurements and associated clinical variables from the MCC20320 cohort are available from the corresponding author upon reasonable request, subject to applicable institutional approvals and participant consent restrictions. Tissue microarray compartment-level measurements and the corresponding histological images and QuPath analysis files are available from the corresponding author upon reasonable request.
Public osteosarcoma gene-expression and clinical data were obtained from the Gene Expression Omnibus under accession GSE33382 and from the TARGET-OS project through the National Cancer Institute Genomic Data Commons (https://portal.gdc.cancer.gov/). Annotated single-cell RNA-sequencing objects were obtained from the OsteoCAR atlas through Figshare (https://doi.org/10.6084/m9.figshare.31029559); the present analysis used specimens from GSE152048 and GSE162454.
The RNA-sequencing data generated from cultured osteoblasts and mesenchymal cells have not yet been deposited in a public repository and are available from the corresponding author upon reasonable request. These data will be deposited in the Gene Expression Omnibus, and the accession number will be included in an updated version of the manuscript.

## Acknowledgements

We thank Maria Lagou for performing the osteogenic differentiation of uc-MSCs. This work was supported in part by the National Cancer Institute (R01CA262802), the National Pediatric Cancer Foundation, and the Montefiore Einstein Comprehensive Cancer Center Support Grant (P30CA013330).

The results published here are in whole or part based upon data generated by the Therapeutically Applicable Research to Generate Effective Treatments (TARGET) initiative, phs000218. The data used for this analysis are available at the Genomic Data Commons (https://portal.gdc.cancer.gov).

---

**Figure S1:**
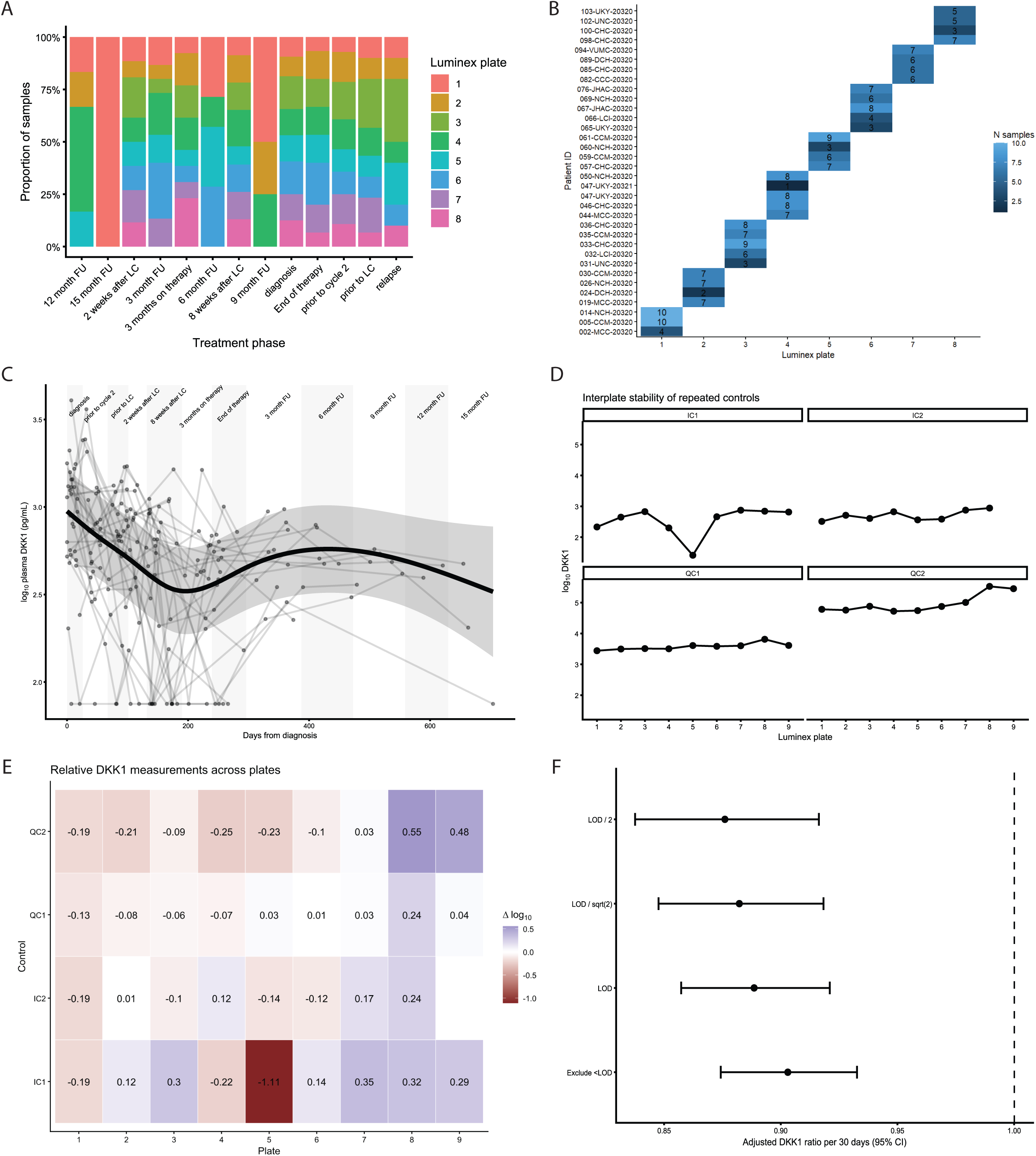
**A)** Distribution of treatment phases across testing plates. **B)** Distribution of patients across testing plates. **C)** Generalized additive mixed model (GAMM) fit for full patient trajectories. Collection stages are herein defined as midpoints between the median timing of consecutive visit stages and are for visualization purposes only. **D)** Interplate comparisons of DKK1 measurements for internal controls (IC) and quality controls (QC) per plate. **E)** Relative DKK1 measurements for internal controls (IC) and quality controls (QC), centred by mean control measurement. **F)** Sensitivity analyses for handling of values below the limit-of-detection (LOD). Comparison of the LMM fit using LOD/2, LOD/√2, full LOD substitution, or exclusion of values below the LOD with 95% confidence intervals.

**Figure S2:**
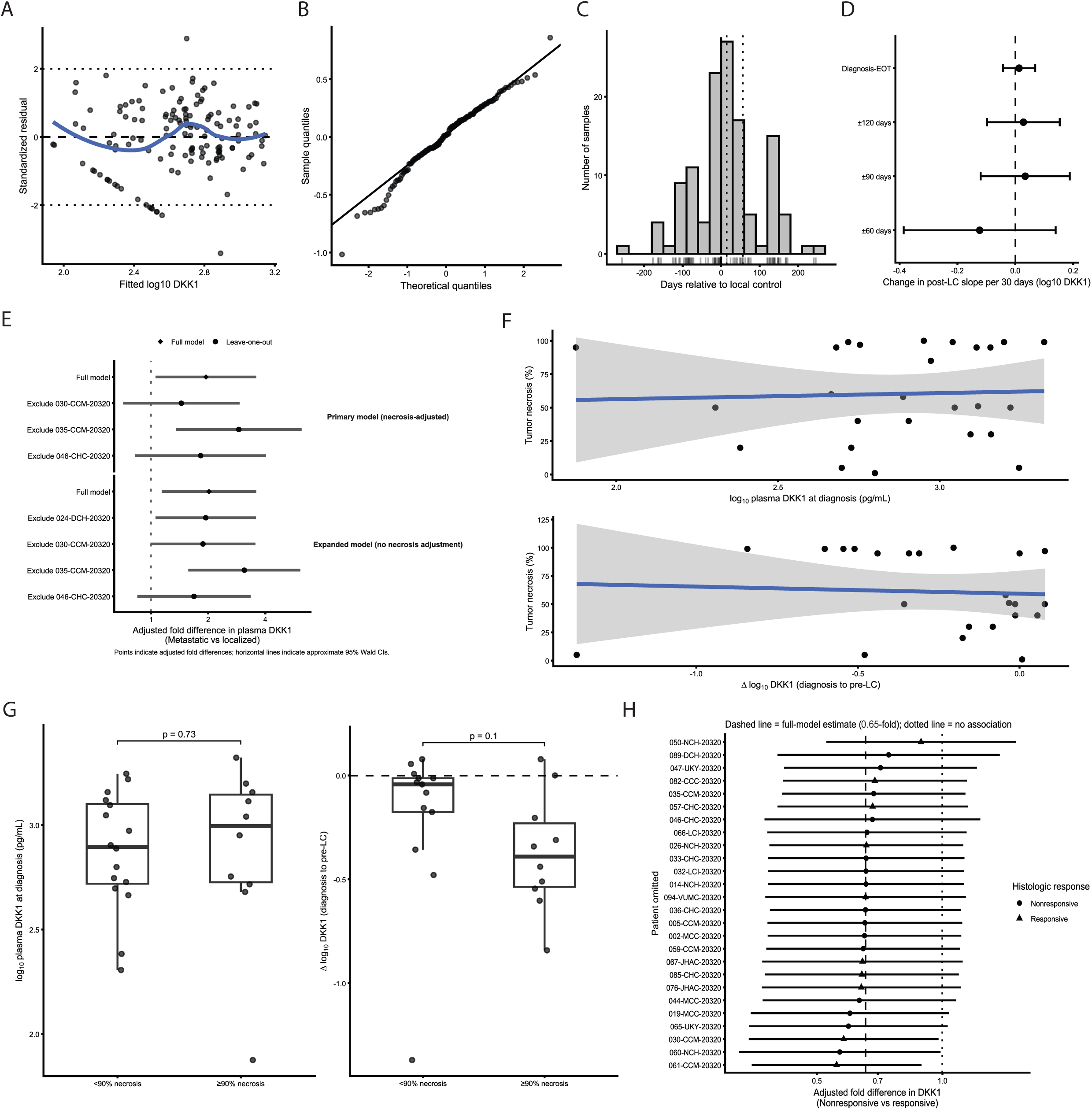
Mixed-effects model diagnostics and sensitivity analyses. **A)** Standardized residuals plotted against fitted values for the final diagnosis-to-end-of-therapy LMM. The blue line shows a LOESS fit. **B)** Q-Q plot of conditional residuals from the same LMM. **C)** Distribution of plasma samples according to days relative to local control. Vertical reference lines denote estimated local control and the first post-local-control collection times. **D)** Sensitivity analysis of the estimated post-local-control slope-change term using the full diagnosis-to-end-of-therapy interval and analyses restricted to ±120, ±90, or ±60 days around local control. Points and horizontal lines indicate estimates and 95% confidence intervals. **E)** Leave-one-metastatic-patient-out sensitivity analyses for the adjusted association between metastatic disease at diagnosis and plasma DKK1 in the necrosis-adjusted model (n = 26) and the expanded model (n = 32) without necrosis adjustment. Diamonds indicate full-model estimates and circles indicate leave-one-out estimates; horizontal lines show approximate 95% Wald confidence intervals. **F)** Tumour necrosis percentage plotted against log10-transformed plasma DKK1 at diagnosis (top) and change in log10-transformed plasma DKK1 from diagnosis to the pre-local-control measurement (bottom). Lines and shaded regions show fitted trends and 95% confidence intervals. **G)** Log10-transformed plasma DKK1 at diagnosis (left) and diagnosis-to-pre-local-control change in log10-transformed DKK1 (right) according to histologic-response group. Points represent individual patients; boxplots show the median and interquartile range; brackets denote Wilcoxon rank-sum comparisons. **H)** Leave-one-patient-out sensitivity analysis of the histologic-response coefficient from the final diagnosis-to-end-of-therapy LMM. Symbols identify the histologic-response group of the omitted patient; horizontal lines indicate approximate 95% Wald confidence intervals. The dashed vertical line indicates the full-model estimate and the dotted vertical line indicates a ratio of 1.

**Figure S3:**
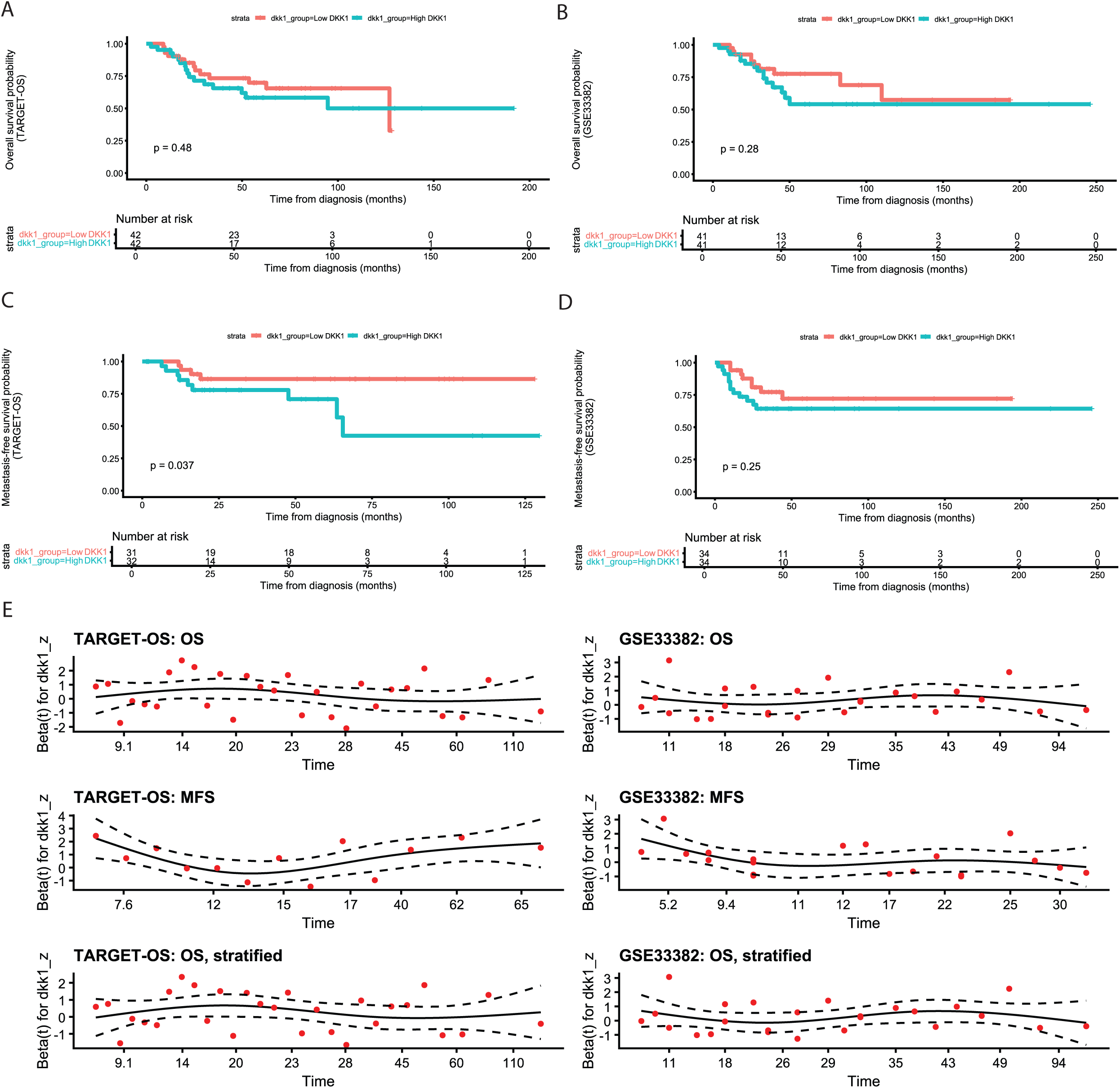
**A)** Overall survival in TARGET-OS, according to median tumour DKK1 expression. **B)** Overall survival in GSE33382, according to median tumour DKK1 expression. **C)** Metastasis-free survival in TARGET-OS, according to median tumour DKK1 expression. **D)** Metastasis-free survival in GSE33382, according to median tumour DKK1 expression. **E)** Scaled Schoenfeld residual diagnostics for the DKK1 coefficient in the continuous Cox models for OS and MFS and in OS models stratified by metastatic status at diagnosis.

**Figure S4:**
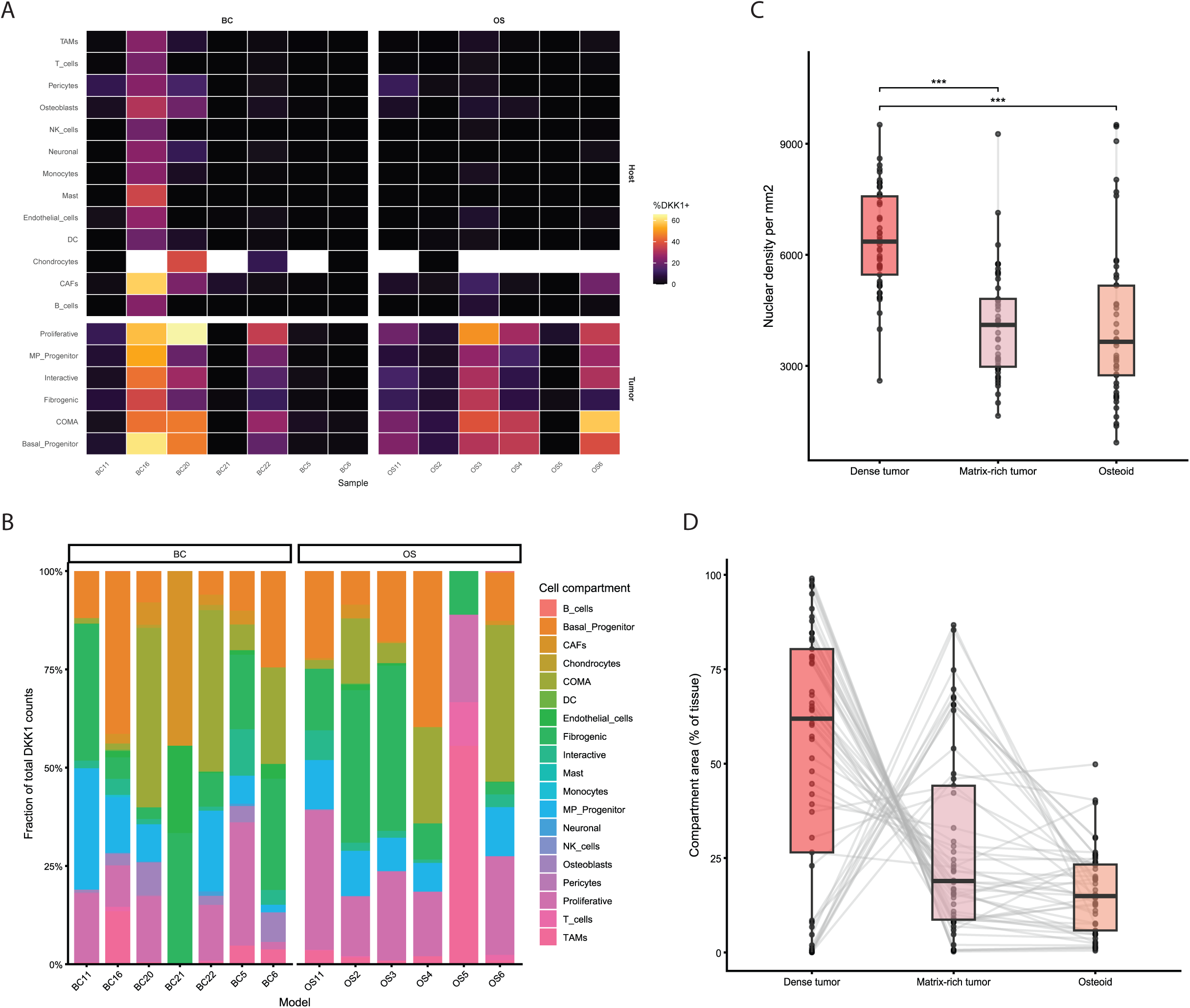
**A)** Fraction of DKK1+ cells by OsteoCAR Level 2 annotation across individual study samples. **B)** Fraction of total DKK1 pseudobulk counts per OsteoCAR Level 2 annotation across study samples. **C)** Nuclear density (n of nuclei per mm²) by QuPath classifier compartment. **D)** Compartment area fraction per TMA core by QuPath classifier compartment. Lines connect compartments from the same TMA core.

## References

1. Luetke, A., et al., Osteosarcoma treatment -where do we stand? A state of the art review. Cancer Treat Rev, 2014. 40(4): p. 523–32.

2. Marina, N.M., et al., Comparison of MAPIE versus MAP in patients with a poor response to preoperative chemotherapy for newly diagnosed high-grade osteosarcoma (EURAMOS-1): an open-label, international, randomised controlled trial. Lancet Oncol, 2016. 17(10): p. 1396–1408.

3. Lagmay, J.P., et al., Outcome of Patients With Recurrent Osteosarcoma Enrolled in Seven Phase II Trials Through Children’s Cancer Group, Pediatric Oncology Group, and Children’s Oncology Group: Learning From the Past to Move Forward. J Clin Oncol, 2016. 34(25): p. 3031–8.

4. Bourhis, E., et al., Wnt antagonists bind through a short peptide to the first beta-propeller domain of LRP5/6. Structure, 2011. 19(10): p. 1433–42.

5. Mao, B., et al., Kremen proteins are Dickkopf receptors that regulate Wnt/beta-catenin signalling. Nature, 2002. 417(6889): p. 664-7.

6. Chen, H., et al., DKK1 Activates the PI3K/AKT Pathway via CKAP4 to Balance the Inhibitory Effect on Wnt/beta-Catenin Signaling and Regulates Wnt3a-Induced MSC Migration. Stem Cells, 2024. 42(6): p. 567–579.

7. Qiang, Y.W., et al., Dkk1-induced inhibition of Wnt signaling in osteoblast differentiation is an underlying mechanism of bone loss in multiple myeloma. Bone, 2008. 42(4): p. 669–80.

8. Gustafson, B. and U. Smith, The WNT inhibitor Dickkopf 1 and bone morphogenetic protein 4 rescue adipogenesis in hypertrophic obesity in humans. Diabetes, 2012. 61(5): p. 1217–24.

9. Zhang, C., H. Dai, and B. de Crombrugghe, Characterization of Dkk1 gene regulation by the osteoblast-specific transcription factor Osx. Biochem Biophys Res Commun, 2012. 420(4): p. 782–6.

10. Krause, U., et al., An unexpected role for a Wnt-inhibitor: Dickkopf-1 triggers a novel cancer survival mechanism through modulation of aldehyde-dehydrogenase-1 activity. Cell Death Dis, 2014. 5(2): p. e1093.

11. Haskell, A., et al., Antisense mediated blockade of Dickkopf 1 attenuates tumor survival, metastases and bone damage in experimental osteosarcoma. Sci Rep, 2025. 15(1): p. 1878.

12. Goldstein, S.D., et al., A monoclonal antibody against the Wnt signaling inhibitor dickkopf-1 inhibits osteosarcoma metastasis in a preclinical model. Oncotarget, 2016. 7(16): p. 21114–23.

13. Tal, A., et al., Inhibition of DKK-1 by WAY262611 Inhibits Osteosarcoma Metastasis. Mol Cancer Ther, 2025. 24(5): p. 728–739.

14. Wang, L., M. Gao, and Z. Liang, Single-cell transcriptomic analysis reveals fatty acid metabolic heterogeneity and identifies DKK1 as a candidate prognostic factor in osteosarcoma. Front Endocrinol (Lausanne), 2026. 17: p. 1918915.

15. Lee, N., et al., A potential role for Dkk-1 in the pathogenesis of osteosarcoma predicts novel diagnostic and treatment strategies. Br J Cancer, 2007. 97(11): p. 1552–9.

16. Ueland, T., et al., Dickkopf-1 enhances inflammatory interaction between platelets and endothelial cells and shows increased expression in atherosclerosis. Arterioscler Thromb Vasc Biol, 2009. 29(8): p. 1228–34.

17. Lee, S., et al., Stroma-derived Dickkopf-1 contributes to the suppression of NK cell cytotoxicity in breast cancer. Nat Commun, 2025. 16(1): p. 1183.

18. Kasoha, M., et al., Dickkopf-1 (Dkk1) protein expression in breast cancer with special reference to bone metastases. Clin Exp Metastasis, 2018. 35(8): p. 763–775.

19. Kuijjer, M.L., et al., Identification of osteosarcoma driver genes by integrative analysis of copy number and gene expression data. Genes Chromosomes Cancer, 2012. 51(7): p. 696–706.

20. National Cancer Institute, Therapeutically Applicable Research to Generate Effective Treatments: Osteosarcoma (TARGET-OS). NCI Genomic Data Commons.

21. Budhathoki, Y., et al., Integrative Single-cell and Spatial Transcriptomic Analysis of Osteosarcoma Reveals Conserved and Distinct Ecosystems Across Sites and Species. bioRxiv, 2026.

22. Voorzanger-Rousselot, N., et al., Platelet is a major contributor to circulating levels of Dickkopf-1: clinical implications in patients with multiple myeloma. Br J Haematol, 2009. 145(2): p. 264–6.

23. Pan, S., et al., Morpholino-driven blockade of Dkk-1 in osteosarcoma inhibits bone damage and tumour expansion by multiple mechanisms. Br J Cancer, 2022. 127(1): p. 43–55.

24. Malladi, S., et al., Metastatic Latency and Immune Evasion through Autocrine Inhibition of WNT. Cell, 2016. 165(1): p. 45–60.

25. Truong, D.D., et al., Mapping the Single-Cell Differentiation Landscape of Osteosarcoma. Clin Cancer Res, 2024. 30(15): p. 3259–3272.

26. Lopez-Fuentes, E., et al., Epigenetic and Transcriptional Programs Define Osteosarcoma Subtypes and Establish Targetable Vulnerabilities. Cancer Discov, 2026. 16(2): p. 296–319.

27. Cai, Y., et al., Inactive Wnt/beta-catenin pathway in conventional high-grade osteosarcoma. J Pathol, 2010. 220(1): p. 24–33.

28. Kansara, M., et al., Wnt inhibitory factor 1 is epigenetically silenced in human osteosarcoma, and targeted disruption accelerates osteosarcomagenesis in mice. J Clin Invest, 2009. 119(4): p. 837–51.

29. Ruiz-Gomez, G., et al., Rational engineering of glycosaminoglycan-based Dickkopf-1 scavengers to improve bone regeneration. Biomaterials, 2023. 297: p. 122105.

30. Moquin-Beaudry, G., et al., Spatial transcriptomic atlas of aggressive osteosarcomas reveals shared immune landscape and targetable surface markers. Nat Commun, 2026. 17(1).

31. Yao, J.C., et al., TGF-beta signaling in myeloproliferative neoplasms contributes to myelofibrosis without disrupting the hematopoietic niche. J Clin Invest, 2022. 132(11).

32. Cao, H., et al., Inhibition of Wnt/beta-catenin signaling suppresses myofibroblast differentiation of lung resident mesenchymal stem cells and pulmonary fibrosis. Sci Rep, 2018. 8(1): p. 13644.

33. Walker, N., et al., Resident tissue-specific mesenchymal progenitor cells contribute to fibrogenesis in human lung allografts. Am J Pathol, 2011. 178(6): p. 2461–9.

34. Reinecke, J.B., et al., Aberrant Activation of Wound-Healing Programs within the Metastatic Niche Facilitates Lung Colonization by Osteosarcoma Cells. Clin Cancer Res, 2025. 31(2): p. 414–429.

35. Kimura, H., et al., CKAP4 is a Dickkopf1 receptor and is involved in tumor progression. J Clin Invest, 2016. 126(7): p. 2689–705.

36. Hayashi, M., et al., Highly personalized detection of minimal Ewing sarcoma disease burden from plasma tumor DNA. Cancer, 2016. 122(19): p. 3015–23.

37. Then, G., Prolactin induction of Amyloid Deposition in the Prostate: A Mechanism for Progression from Benign Prostatic Hyperplasia to Prostate Cancer. 2026, CUNY Lehman College: CUNY Academic Works.

38. San Martin, R., et al., Tenascin-C and Integrin alpha9 Mediate Interactions of Prostate Cancer with the Bone Microenvironment. Cancer Res, 2017. 77(21): p. 5977–5988.

39. San Martin, R., et al., Transcriptional profiling of Hutchinson-Gilford Progeria syndrome fibroblasts reveals deficits in mesenchymal stem cell commitment to differentiation related to early events in endochondral ossification. Elife, 2022. 11.

